# Is Your Style Transfer Doing Anything Useful? An Investigation Into Hippocampus Segmentation and the Role of Preprocessing

**DOI:** 10.1101/2024.08.22.24312425

**Authors:** Hoda Kalabizadeh, Ludovica Griffanti, Pak-Hei Yeung, Natalie Voets, Grace Gillis, Clare Mackay, Ana IL Namburete, Nicola K Dinsdale, Konstantinos Kamnitsas

## Abstract

Brain atrophy assessment in MRI, particularly of the hippocampus, is commonly used to support diagnosis and monitoring of dementia. Consequently, there is a demand for accurate automated hippocampus quantification. Most existing segmentation methods have been developed and validated on research datasets and, therefore, may not be appropriate for clinical MR images and populations, leading to potential gaps between dementia research and clinical practice. In this study, we investigated the performance of segmentation models trained on research data that were *style-transferred* to resemble clinical scans. Our results highlighted the importance of intensity normalisation methods in MRI segmentation, and their relation to domain shift and style-transfer. We found that whilst normalising intensity based on min and max values, commonly used in generative MR harmonisation methods, may *create* a need for style transfer, Z-score normalisation effectively maintains style consistency, and optimises performance. Moreover, we show for our datasets spatial augmentations are more beneficial than style harmonisation. Thus, emphasising robust normalisation techniques and spatial augmentation significantly improves MRI hippocampus segmentation.

## 1 Introduction

Many neurodegenerative diseases cause volumetric atrophy in the region of the hippocampus [1], including Alzheimer’s disease (AD). AD is clinically characterised by a progressive decline in cognitive function with diagnosis and monitoring of the disease commonly including the assessment of hippocampal atrophy in brain MRI scans [2]. Specifically, the volume of the hippocampus, is often measured, either through manual or automated segmentation.

Manual segmentation requires large amounts of time and expert knowledge and suffers from inter-rater variability. Therefore, there is a demand for accurate automated hippocampus segmentation methods [3]. Among different types of techniques, deep learning (DL) based methods show great promise for the segmentation of the hippocampus, outperforming traditional atlas-based approaches [4], [5]. However, training CNNs generally requires the availability of manual labels, limiting the applicability in clinical practice. Furthermore, due to differences in image acquisitions and patient demographics, models trained on research datasets are unlikely to generalise to clinical populations. Therefore, there is a need to overcome this *domain shift* between the source (research) and target (clinical) dataset, enabling the development of segmentation models for clinical scans without requiring segmentation labels.

To address general domain shift, data augmentation is a commonly used technique for artificially enhancing the diversity of training data, to increase model generalisability and robustness. Augmentation has been shown to improve down-stream segmentation performance across brain imaging studies [6]. However, augmentation requires the identification and modelling of differences between domains, which is non-trivial in the presence of varying populations and scanner technologies. Another related field of research is image-to-image (I2I) translation, which is an approach that aims to learn the mapping between different visual domains, mostly based on generative models. For instance, Pix2pix [7] utilises a conditional GAN to map between image domains, but relies on pixel-to-pixel correspondence, limiting its applicability to MR images from different sites. CycleGAN [8] overcomes the need for paired data using cycle consistency. MR harmonisation approaches, e.g., [9] are based on I2I methods, aiming to overcome the style-based domain shifts associated with differing acquisition scanners while maintaining the underlying anatomy.

Therefore, in this study, we aim to investigate which techniques are effective for overcoming the *domain shift* between our source (research) dataset and target (clinical) dataset for the task of MR hippocampus segmentation. Our contributions are as follow:

— We demonstrate the use of a 2-stage pipeline for generating style-transferred images that are subsequently used to train a hippocampus segmentation model.
— We explore the impact on downstream hippocampus segmentation performance of different preprocessing and augmentation approaches.
— We show that the use of appropriate normalisation (*i*.*e*. Z-score normalisation) and spatial augmentation (*i*.*e*. paired affine registration) can lead to substantial improvements on downstream hippocampus segmentation performance, even without a sophisticated style transfer pipeline.

The findings of this study may shed light on the importance of developing a robust preprocessing pipeline for MR hippocampus segmentation in future studies.

## 2 Methods

To overcome the domain shift between the research and clinical data, we implemented a 2-stage approach, formed of a style transfer (ST) network followed by a segmentation network. A schematic of this pipeline is shown in Figure 1. We assumed access to a source (research) dataset, *𝒟*_*s*_ = *{****X***_*s*_, ***Y***_*s*_*}*, and an unlabelled target (clinical) dataset, *𝒟*_*t*_ = *{****X***_*t*_*}*, to first train a style transfer model that generates source images in the style of target images, 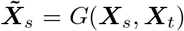, following which we used the ST images to train a segmentation model 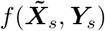, such that the performance for *𝒟*_*t*_ is maximised.

**Fig. 1:**
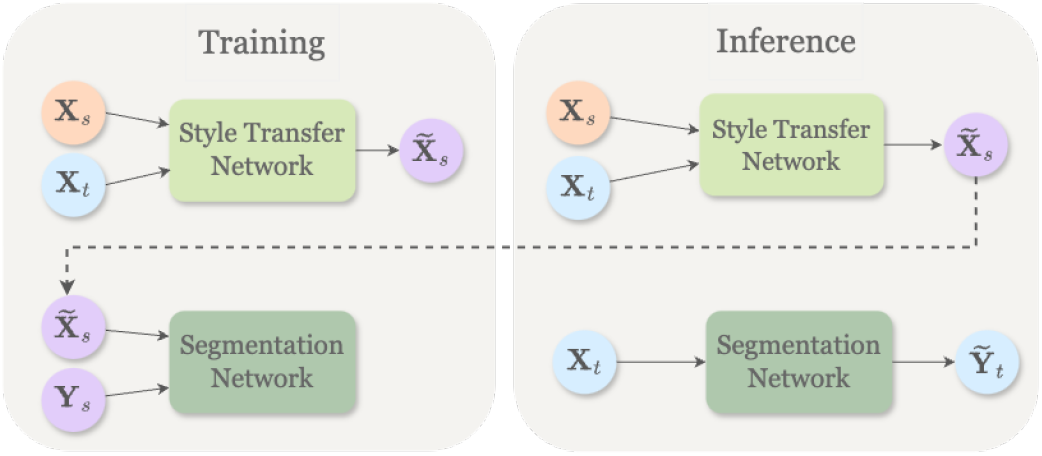
A schematic of the proposed 2-stage pipeline.

### 2.1 Style Transfer: Style-Encoding GAN

We utilised the Style-Encoding GAN (SE-GAN) [9] for our style transfer network. Similarly to StarGANv2 [10], it is formed of a single generator (G), discriminator (D), mapping network (M) and a style encoder (E). During training, SE-GAN trains G to generate diverse images corresponding to a single image slice ***x*** *∈* ***X*** using a style code *c*, provided by either M or E. Consequently, the generator *G* translates an input image, ***x***, into an output image, 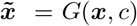, that is reflective of the style of *c*. To validate the successful injection of *c* into the output image 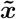, *E* is used to extract the style code from images. The style code is a 1 *×* 64 vector, allowing *E* to produce diverse style codes from different images. Moreover, the discriminator *D* learns to classify images as real or fake, as produced by *G*(***x***, *c*). In our experiments, *G* is used to synthesise output images 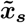 based on source images ***x***_*s*_ that are reflecting the style *c* extracted from various reference images in ***X***_*t*_. Finally, 3D volumes can be reconstructed by stacking the 2D slices. The network is trained using the loss introduced in [10], formed of an adversarial loss *L*_*GAN*_, cycle consistency loss *L*_*cyc*_, style reconstruction loss *L*_*sty*_ and diversification loss *L*_*div*_, weighted by *λ*_*cyc*_, *λ*_*sty*_ and *λ*_*div*_ respectively, resulting in the following objective function:

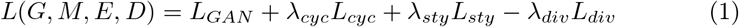

### 2.2 Segmentation: U-Net

The second stage of the framework is the training of the segmentation network, *f*, for which we used a 3D U-Net [11]. The network is trained with a Dice loss, *L*_*dice*_, using the style transformed source data such that:

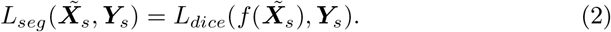

### 2.3 Investigating Preprocessing

#### Registration

To mitigate content shift, defined as variations in anatomical alignment between brain scans, we investigated the impact of registration on both style transfer and downstream segmentation performance. To this end, we conducted registration, using both 6 (rigid-body) and 12 (affine) degrees of freedom (DOF) and compared two main approaches. (1) **MNI-Reg**: A spatial harmonisation approach, where both source and target images are registered to a standard space, (2) **Paired-Reg**: A spatial augmentation approach, where every source image is registered to every target image.

#### Normalisation

In most ST models intensity normalisation is performed during training, through linear scaling of the range of intensities from [Min, Max] to a pre-defined range such as [0,1], which we call Min-Max normalisation. This approach, however, is often unsuitable for MRI images as they can have varying intensity distributions from different acquisitions, leading to inconsistent normalisation results. Additionally, intensity outliers common in MRI data can skew normalisation. We investigated the performance of Min-Max normalisation and explored the impact of applying Z-score normalisation on a per-subject basis.

## 3 Experimental Setup

### 3.1 Datasets

#### Research Dataset

The HarP dataset [12] was used as the labelled *research dataset*, consisting of 135 T1-weighted MRI volumes (cognitively normal controls, MCI and AD patients) from a range of scanners, and corresponding hippocampus masks [13]. All MRIs were registered to MNI-space.

#### Clinical Dataset

We used a dataset from the Oxford Brain Health Clinic (OBHC) [14] as our *clinical dataset*, representing the unlabelled target domain. It includes 29 patients referred to a memory clinic, who agreed to the use of data for research. The lack of strict inclusion criteria typical of a dementia research study, makes this dataset representative of real-world memory clinic patients. The scans were collected using a 3T Siemens scanner. Hippocampi were manually annotated by a clinician. BHC labels were used only for model evaluation, not for training.

Figure 2 compares the hippocampal volumes between the research (HarP) and clinical (BHC) populations. It can be seen that generally the research population have larger hippocampal volumes than the clinical group. This difference can probably be attributed to dementia research typically recruiting patients that tend to be younger and have less hippocampal atrophy [15].

**Fig. 2:**
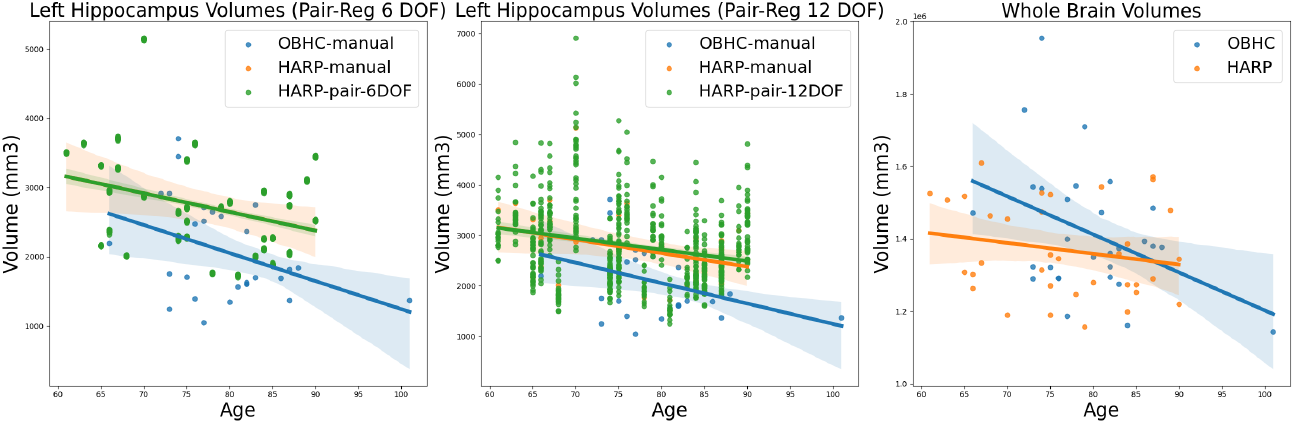
Volume against age plots for HarP and OBHC: left hippocampus volumes for Paired-Reg 6 DOF (left), 12 DOF (middle), and whole brain volumes (right).

### 3.2 Preprocessing

For anonymisation, the BHC scans were brain extracted and thus we performed brain extraction on the HarP dataset. N4 bias field correction was used to correct for low-frequency intensity non-uniformity. Images were split into left and right hemispheres for training. Data registration followed Section 2.3.

### 3.3 Implementation Details

For training the ST network, 132 HarP images were used as the source images, and 20 randomly selected BHC images were used as the references, using a learning rate of 10^*−*4^ and the Adam optimiser. For Equation 1, we set *λ*_*cyc*_ = 10, *λ*_*sty*_ = 1 and *λ*_*div*_ = 1, as suggested by [9]. Moreover, 100 HarP images were used for training and then the trained ST network was used to generate a style-transferred image for each HarP image (N=32) in the style of each BHC image (N=20) resulting in 640 style-transferred images, which were used to train the segmentation model.

The chosen U-Net architecture network had four downsampling and upsampling layers, whereby each layer was formed of a convolutional layer, a ReLU activation function and a batch normalisation layer. The depth, defined as the number of convolutions, doubled between each layer, starting with 4. The U-Nets were trained using a learning rate of 10^*−*3^, and the Adam optimiser. The training was conducted using 3-fold cross-validation and tested on 9 BHC patients (18 hippocampi) that were not used during the training or validation. Training was conducted using an Nvidia A10 GPU. The ST and segmentation networks required an average training time of 35 hours and 12 hours, respectively. However, once training was complete, the segmentation model’s testing, or inference, took only a few seconds per scan, making it suitable for clinical applications.

## 4 Results & Discussion

### 4.1 Domain Shift

First, to establish the domain shift between the datasets, we tested publicly available out-of-the-box (OOB) tools: FSL FIRST [16], FreeSurfer [17], SynthSeg [5], Hippodeep [18], as well as U-Net based approaches: a U-Net trained solely on HarP, basic augmentation (affine transforms, flips, noise, intensity changes), MRI-specific augmentation (motion, bias field). We also compared with adversarial unsupervised domain adaptation, approach shown potent for tackling domain shift in medical imaging [19], and specifically the model developed in [20] (UDA).

Table 1 shows the dice score (DSC) values for the different approaches. The OOB models all performed better on our research population compared to our clinical population, achieving maximum DSC of 0.829 and 0.758, respectively. In particular, FreeSurfer and FIRST had instances of complete failure for the OBHC data (DSC = 0). For most patients, UDA was comparable to the augmentation methods, however, when examining the worst-case scenarios, UDA led to particularly low dice scores for certain individuals. Data augmentation, thus, proved to be the most effective approach for performance enhancement. These findings demonstrate the limitations of existing methods and highlight the potential value of exploring more sophisticated data augmentation approaches.

**Table 1:**
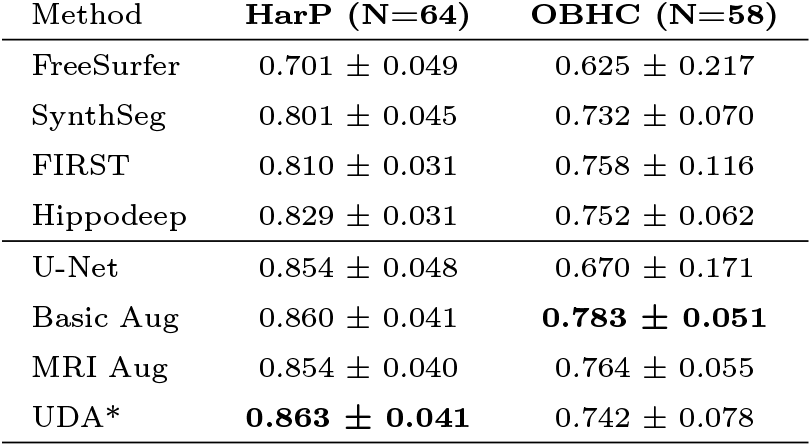
DSC for segmentation methods on HarP (Source) and OBHC (Target). N is the number of test hippocampi (i.e., 2*×* number of patients). * UDA test sizes were smaller due to training on a sample of unlabelled OBHC.

### 4.2 Registration

We then explored the effect of the choice of registration approach. Figure 2 shows the effect of registration on the hippocampal volumes: as rigid registration only involves translation and rotation for brain alignment, there is no change between the original HarP volumes and those registered to OBHC. However, affine registration performs global scaling, resulting in a slight increase in volume of the registered HarP hippocampi. Although the OBHC registration targets have distinctly smaller hippocampal volumes, they have, on average, larger whole-brain volumes, leading to larger hippocampi in the registered HARP images.

### 4.3 Style Transfer vs Normalisation

Table 2 shows the segmentation performance for models trained on Z-score and Min-Max normalised data, and tested on the 9 unseen OBHC patients (18 hippocampi), for a range of evaluation metrics, namely the Dice score (DSC), Hausdorff distance (HD), and Relative Absolute Volume Difference (RAVD). As a supervised benchmark, we trained directly on the OBHC dataset (20 labelled patients), which achieved an average DSC of 0.744. By comparison, simply training on the Min-Max normalised HarP dataset achieved a mean DSC of 0.616, clearly indicating presence of a domain shift. The results of training with the U-Net on the HarP dataset with the different registration schemes, normalisation schemes and the use of ST can then be seen. MNI-Reg-6 ST led to a 4% increase in performance, with a DSC of 0.651. Z-score normalisation outperformed Min-Max normalisation across the experiments. Without Z-score normalisation, a noticeable style shift exists, which can be slightly mitigated by training on style-transferred images (MNI-Reg-6 ST). However, implementing Z-score normalisation effectively reduces this style shift, increasing performance to levels similar to a model trained on target data, while the benefits offered by style transfer are reduced. Following this, the impact of mitigating content shift using affine registration was evaluated, specifically employing the paired registration approach (Table 2). A significant increase in segmentation performance is observed through augmenting the data with paired registration (Paired-Reg-12), achieving an average DSC of 0.780 without ST and 0.787 with. Standard augmentations further improved the performance, achieving the highest DSC of 0.797 (Paired-Reg-12 ST + Aug). The source, reference and style-transferred images, generated by ST networks trained on affine paired registered images (Paired-Reg-12 ST) using both normalisation approaches, have been visualised in Figure 3. The figures reveal that Min-Max normalisation tends to highlight the style transfer effect more visibly than Z-score normalisation. This difference arises because Min-Max normalisation is sensitive to extreme values in MRI data, which can distort the results. In contrast, Z-score normalisation is more robust to such outliers. Thus, the differences between the figures likely reflect variations in intensity ranges rather than style transfer performance. This difference is further demonstrated by the intensity distributions plotted in the Supplementary Material. Figure 4 provides a qualitative comparison between the manual segmentations and the best performing model predictions (Paired-Reg-12 ST with augmentations) on the OBHC test data.

**Table 2:** Segmentation results using different normalisation and registrations. N is the number of train hippocampi (i.e. 2*×* number of patients)

| Train Data | Train N | Norm | Min DSC ↑ | Avg DSC ↑ | 95 % HD ↓ | RAVD↓ |
| --- | --- | --- | --- | --- | --- | --- |
| OBHC | 40 | Z-score | 0.617 | 0.744±0.016 | 3.865±1.357 | 18.388±1.694 |
| HarP | 64 | Min-Max | 0.480 | 0.616±0.037 | 5.264±0.970 | 97.972±9.456 |
| MNI-Reg-6 ST | 1,280 | Min-Max | 0.521 | 0.651±0.015 | 4.036±0.06 | 83.033±2.037 |
| HarP | 64 | Z-score | 0.674 | 0.746±0.009 | 6.013±1.297 | 26.326±1.856 |
| MNI-Reg-6 ST | 1,280 | Z-score | 0.688 | 0.757±0.008 | 5.072±1.391 | 20.736±1.906 |
| Paired-Reg-12 | 1,280 | Z-score | 0.703 | 0.780±0.005 | 3.452±0.237 | 20.009±0.913 |
| Paired-Reg-12 ST | 1,280 | Z-score | 0.672 | 0.787±0.004 | 3.169±0.250 | 23.794±4.033 |
| Paired-Reg-12 ST<br>+ Aug | 1,280 | Z-score | 0.719 | 0.797±0.003 | 3.133±0.143 | 27.509±3.724 |

**Fig. 3:**
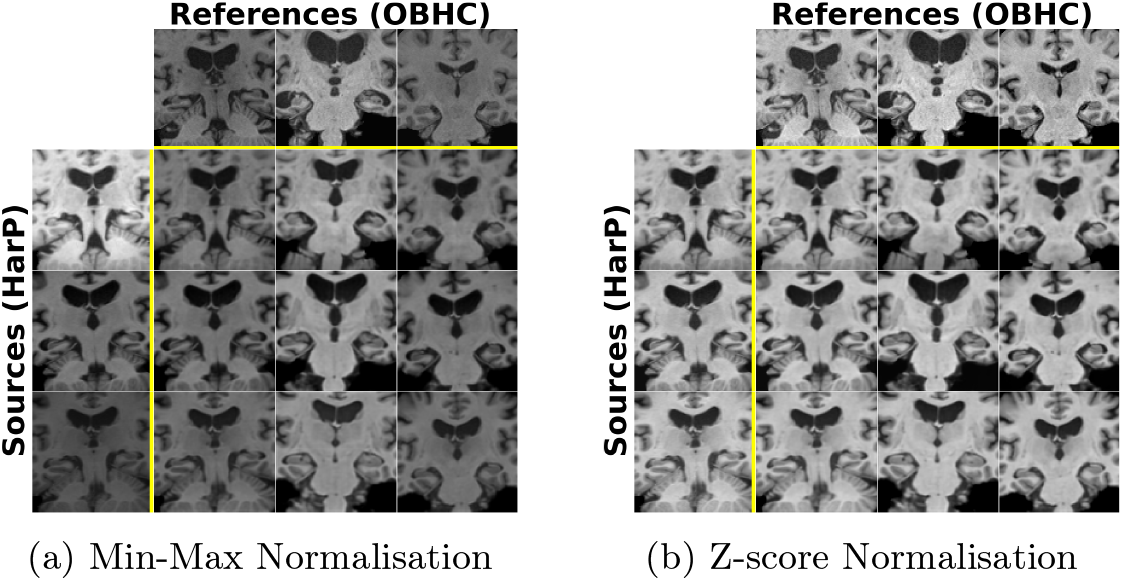
Generated ST images for a given source (first column) and reference (first row), using a) Min-Max and b) Z-score normalisation.

**Fig. 4:**
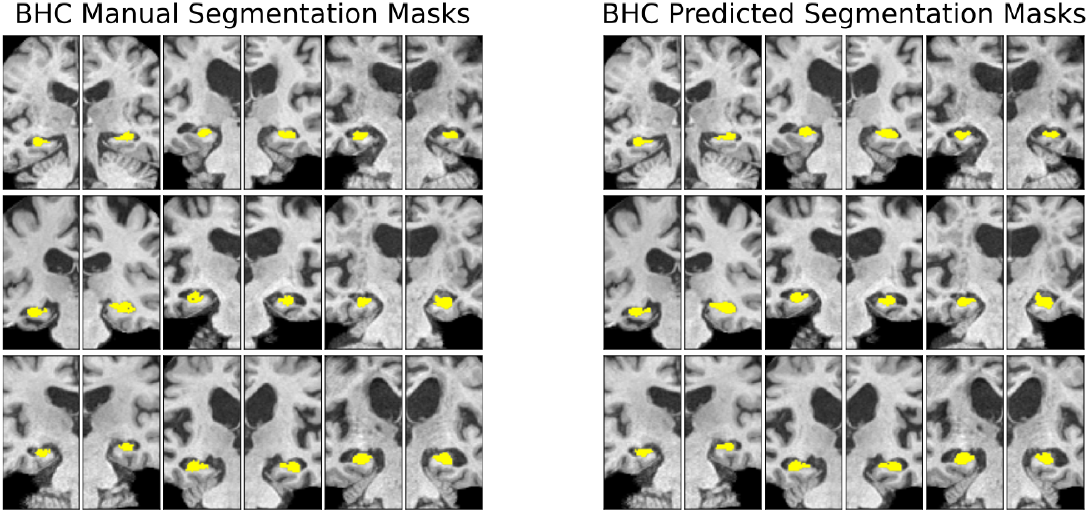
Manual and predicted segmentation masks for the OBHC test data.

## 5 Conclusion

In conclusion, we implemented a 2-stage pipeline consisting of a ST and a segmentation network. Our experimental findings underscored the significance of normalisation methods in MRI augmentation and segmentation tasks. While experiments with Min-Max normalisation may suggest a style shift and the potential benefits of style transfer, this interpretation may be misleading and is a result of inappropriate normalisation. Our findings indicate that Z-score normalisation negates the necessity for style transfer by effectively maintaining style consistency in MRI data, thereby optimising segmentation performance directly. Moreover, for the task of hippocampus segmentation, our results demonstrate that mitigating the content shift using a spatial augmentation approach (i.e. Paired-Reg 12 DOF) can be far more beneficial than a spatial harmonisation approach, such as aligning all images to MNI. The improved performance may be attributed to the spatial diversity introduced by the augmentation, which enhances segmentation robustness. Thus, prioritising robust normalisation techniques and appropriate spatial augmentation can lead to substantial improvements in the generalisability of MRI segmentation. Future studies may, thus, benefit from considering spatial augmentation, akin to those currently employed in style transfer, to achieve further improvements in hippocampus segmentation performance.

## Data Availability

HarP dataset is available online. Access to the OHBC dataset can be requested via DPUK.

http://www.hippocampal-protocol.net/SOPs/index.php

https://portal.dementiasplatform.uk/

## Acknowledgments

The authors are grateful for support from: the University of Oxford Department of Computer Science Scholarship (HK), the Bill and Melinda Gates Foundation (NKD, AILN) and the Presidential Postdoctoral Fellowship (Nanyang Technological University) (PHY). We are grateful to the operations team of the OBHC. The OHBC data collection and analysis is supported by the NIHR Oxford Health Biomedical Research Centre (NIHR203316) - a partnership between the University of Oxford and Oxford Health NHS Foundation Trust, the NIHR Oxford Cognitive Health Clinical Research Facility, and the Wellcome Centre for Integrative Neuroimaging (203139/Z/16/Z, 203139/A/16/Z). The views expressed are those of the author(s) and not necessarily those of the NIHR or the Department of Health and Social Care. For the purpose of open access, the authors have applied a CC BY public copyright licence to any Author Accepted Manuscript version arising from this submission.

