## Supplementary Material for "Is Your Style Transfer Doing Anything Useful? An Investigation Into Hippocampus Segmentation and the Role of Preprocessing"

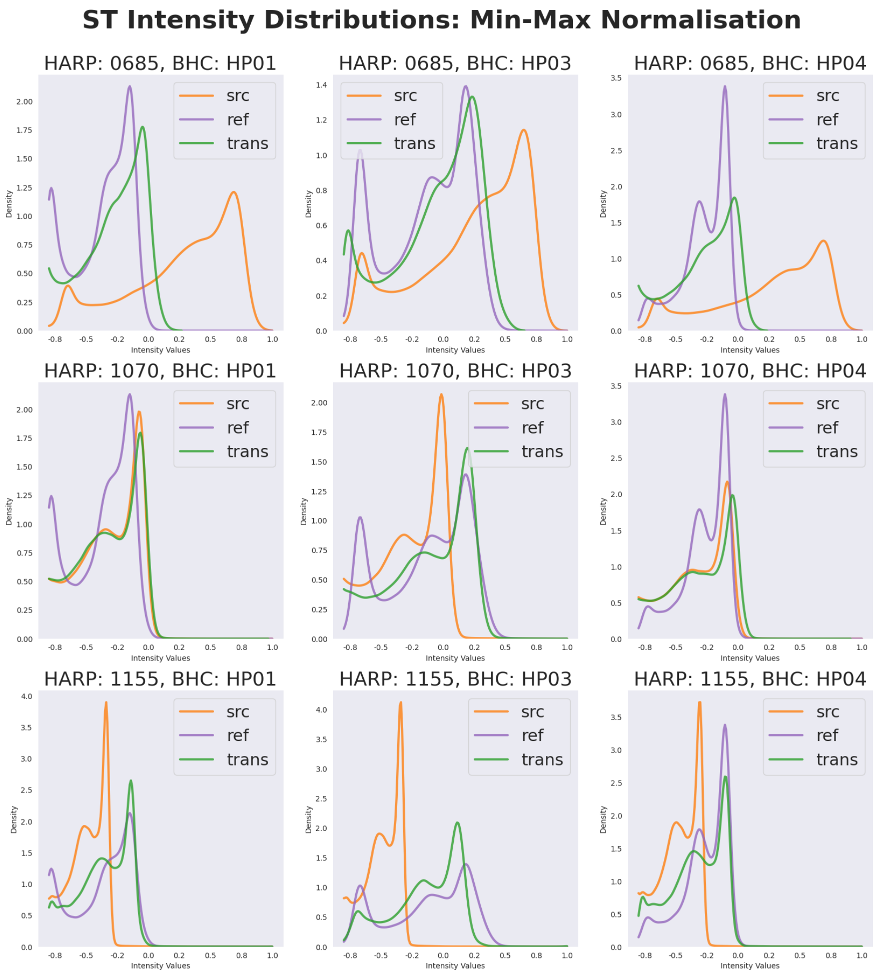


Fig. 1: Individual plots of intensity distributions. Each figure shows the distributions of the following scans: source (orange), reference (purple) and Paired-Reg-12 Style Transformed (green). All scans have been normalised using Min-Max normalisation.

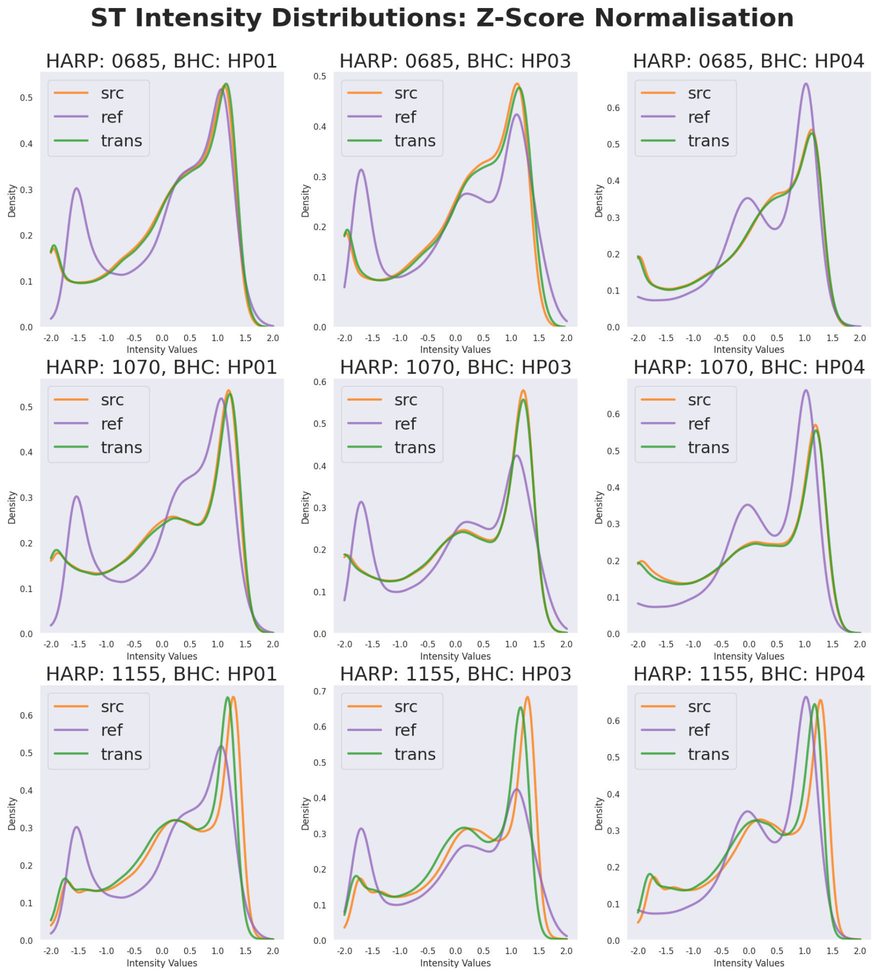


Fig. 2: Individual plots of intensity distributions. Each figure shows the distributions of the following scans: source (orange), reference (purple) and Paired-Reg-12 Style Transformed (green). All scans have been normalised using Z-score normalisation.
